# Machine Learning for Neonatal Mortality Risk Assessment: A Case Study Using Public Health Data from São Paulo

**DOI:** 10.1101/2020.05.25.20112896

**Authors:** Carlos Eduardo Beluzo, Luciana Correia Alves, Rodrigo Bresan, Natália Arruda, Ricardo Sovat, Tiago Carvalho

## Abstract

Infant mortality is a reflection of a complex combination of biological, socioeconomic and health care factors that require various data sources for a thorough analysis. Consequently, the use of specialized tools and techniques to deal with a large volume of data is extremely helpful. Machine learning has been applied to solve problems from many domains and presents great potential for the proposed problem, which would be an innovation in Brazilian reality. In this paper, an innovative method is proposed to perform a neonatal death risk assessment using computer vision techniques. Using mother, pregnancy care and child at birth features, from a dataset containing neonatal samples from São Paulo city public health data, the proposed method encodes images features and uses a custom convolutional neural network architecture to classification. Experiments show that the method is able to detect death samples with accuracy of 90.61%.

## 1 Introduction

Infant mortality rate (IMR) is one of the most important measures for indicating regional and national public health care status of children. IMR is, therefore, a very relevant index of a country’s condition in terms of health or development. A decrease in IMR results in survival improvements, which can positively influence the national public health indicators.

The first 28 days of life - known as the neonatal period - represent the most vulnerable moment for a child survival and are strongly influenced by unfavorable life conditions of the population and by health care. Hence, mortality during this period reflects the complex conjunction of biological, socioeconomic and care factors, the latter related to attention to the pregnant woman and the newborn [13].

About 46% of the world’s deaths occur among children under five years of age. Most of these deaths are concentrated on the first day and the first week of life. The main causes of neonatal deaths are congenital malformation, asphyxia during labor and perinatal infections; that is, mostly preventable deaths from health services [7].

Mosley and Chen [17] proposed a hierarchical model based on the hypothesis that socioeconomic factors determine behaviors, which, in turn, have an impact on a set of biological factors. According to their model, biological factors are those directly responsible for death. The hierarchical model brings a great advance to the development of public policies, since information coming from studies that are limited to only a group of risk factors result in inadequate recommendations to assess the deaths among children, as they presents a limited vision of the phenomenon [8]. It is not hard to see how paramount is to develop and deploy methods to support policy makers on their strategies to deal with neonatal mortality.

In Brazil, between 1999 and 2013, although the number of infant deaths have declined, 70% of the deaths occurred due to preventable causes. Among the main causes, those related to adequate attention to pregnancy and delivery presented growth of about 33% [21]. Furthermore, neonatal mortality rates (amount of dies before 28 days of live per 1000 life live births) starts to point to a slightly increase trend, specially in 2015 and 2016, in different Brazilian regions, as depicted in Figure 1.

**Fig. 1.**
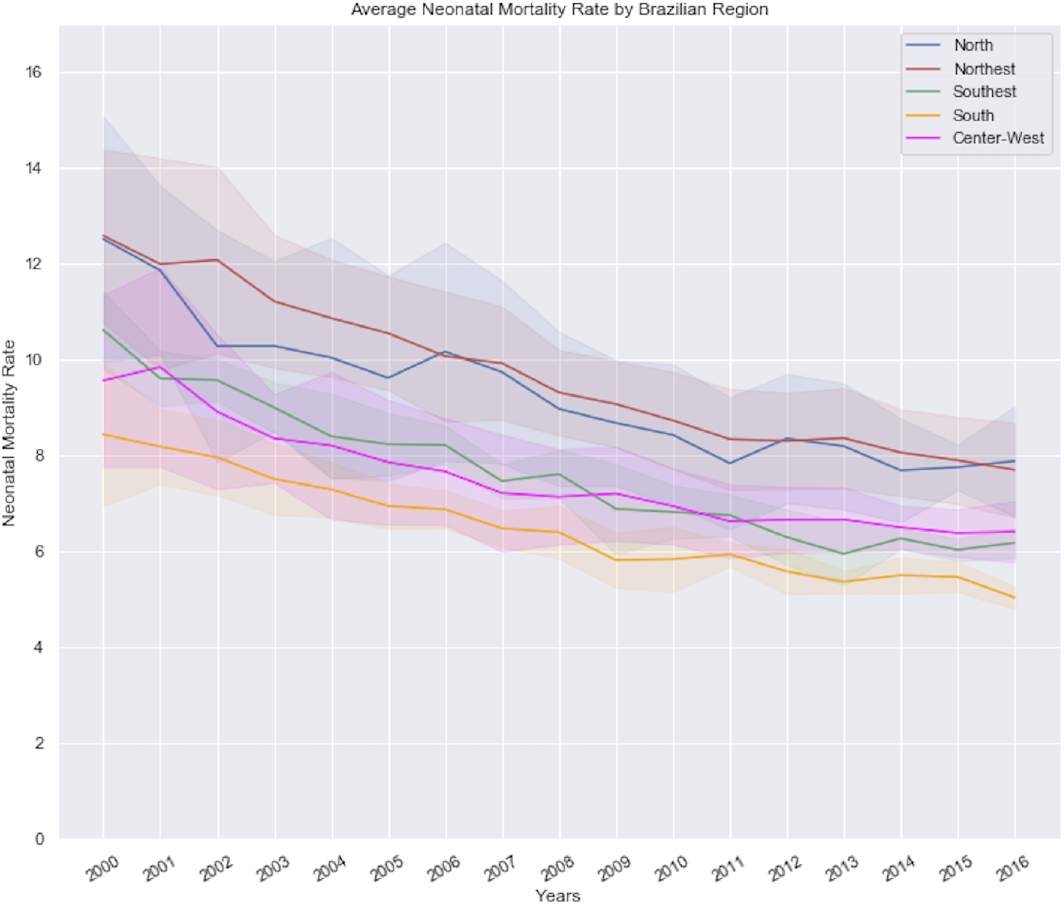
Neonatal mortality rate from 2000 to 2016. Bold lines represent average rate by Brazilian geographical region while shadows represent standard deviation.

Brazilian’s municipalities are responsible for taking care of public health policy focused on infant mortality (IM) [16,2]. Specifically in São Paulo^1^, neonatal mortality rate is the lowest when compared against other municipalities in Brazil.

In parallel to the concerning scenario of Brazilian neonatal mortality trends, application of machine learning (ML) algorithms in public health problems is a very recent research field and a very small number of conducted studies have been proposed. Hsieh *et al*. [12] compared ML models with the objective of predicting mortality in patients undergoing unplanned extubation in intensive care units. The authors observed that even with a limited data set comprising 341 samples, some models obtained significant results. The best results were obtained using a Random Forest model with an unbalanced data set, resulting in an precision of 88% and a recall of 85%.

Pan *et al*. [18] proposed adoption of these models to identify women with high risk pregnancy, an approach to complement the paper question form applied in risk assessment at social services in Illinois. Proposed methods were capable of improving the efficiency in decision making process, improving identification of women with high-risk pregnancy eligible to receive specific health services.

Podda *et al*. [19] uses a Neural Network to estimate the chances of survival of premature newborns. The authors compare their approach with the most common logistical methods. With superior performance, the method proposed by the authors allows better identification of risks and leads to improvements in the quality of the decision-making and risk assessment process. In addition, the authors explain that while logistic regression and other linear models are easily understandable, this ease of interpretation is lost when interactions between variables are present and, in this case, neural networks may work better.

New studies are been conducted focused on the application of ML on problems involving data from the Demography domain. Recently Arruda [3] proposed the use of ML algorithms to establish determinants of adult probability of death. This works investigated the relationships among socioeconomic, structural, contextual and health factors and the adult probability of death, in the some Brazilian micro regions, using data extracted from official Brazilian information systems.

Pan et.al. [18] used ML algorithms to evaluate the positive predictive value for early assessment of adverse birth risk among pregnant women as a means of improving the allocation of social services. Administrative data from Illinois Department of Human Services were used to develop a ML model for adverse birth prediction and improve upon the existing paper-based risk assessment.

Given that neonatal mortality is a complex phenomenon, involving interactions of several characteristics, requiring a large volume of data for its full understanding, the application of ML methods in this context is not only pertinent but also an innovative to Brazilian reality, specially using public health data, and brings substantial contributions and insights to the neonatal mortality problem. At this paper we propose a new method for neonatal mortality risk assessment by projecting categorical features into an image domain and then classifying samples according to death risk, using a ML method.

Computational methods based on image processing and ML have been applied on different domains [4,14], even when data is not originally encoded as images [11,20], achieving expressive results, which suggest that its application on IM problem would bring substantial contributions.

On the method proposed on this paper, neonatal mortality is represented as a binary classification problem and using a new Convolutional Neural Network (CNN) architecture, developed a model to deal with our encoded images and named it DemogNet (Demographic Network). We achieved an accuracy of 90.61% in a dataset with more than 1.4 million samples.

To the best of our knowledge, this is the first time this kind of data/problem have been addressed using a combination of demographic/socioeconomic features, and computer vision techniques. Main contributions of proposed method include: (1) proposition of a new CNN architecture specialized to deal with very small gray scale images; (2) an accuracy of more than 90% in the task of neonatal death risk assessment, in a data set comprising more than 1.4 million samples; (3) a feature importance evaluation, which allows better understanding for the problem of infant mortality.

Rest of this work is divided as follow: Section 2 presets details of the dataset used in the experiments. Section 3 describes proposed method on this paper, including the procedure implemented for encoding tabular data into gray scale images and details about proposed CNN architecture. Section 4 describes performed experiments and results. Finally, Section 5 presents a conclusions and directions for future works.

## 2 Data Sources and Dataset Construction

Proposed method was validated using data extracted from Brazilian Information System of Live Births (SINASC - Sistema de Informações sobre Nascidos Vivos) and from the Brazilian Information System of Mortality (SIM - Sistema de Informações sobre Mortalidade)^2^, to construct a unweighted labeled dataset comprising more then 1.4 millions of images from two classes: death and alive. Specifically, we use data from São Paulo city from 2012 to 2018, resulting in a dataset with 1,435,834 samples. The number of positive class (death) represents 0.6% of total amount of data.

SINASC integrates epidemiological information regarding alive births in Brazil. Its records are associated with new born and its parents, features as birth location, birth date, sex, APGAR score [1], weeks of gestation, mother’s address, age, schooling, occupation, number of children, type of delivery, quantity of prenatal medical appointments, anomalies, and others.

SIM provides features from dead individuals like death main cause^3^, birth date, location, death date, age, sex, race, and many others. Furthermore it also registers data regarding relatives of the person like mother’s age, number of children, gestation weeks, and many others social and economic variables. So, this system is a very important source of information to performs demography studies, including IM.

Only SINASC features, described on Table 1, were used as input features for the method. SIM data are applied just for labelling purpose, so for each SINASC record, SIM data were used to label the sample as death or alive class, making possible to construct a big annotated dataset. After the linkage between SIM and SINASC, the key used on the joining operation were removed from the resultant dataset, as well as many others fields that could be used to re-identify individuals. The column named “FTV” represents values used during the image encoding process, described afterwards.

**Table 1.** TABLE 1 - Dataset features details

| Feature | Features Domain | Details | FTV |
| --- | --- | --- | --- |
| n_tp_ocorrencia | 0-4 | Birth place code | n/a |
| n_nu_idade | 0-63 | Mother Age | n/a |
| n_tp_estado_civil | 0,1,2,3,4,5,9 | Mother marital status | +100 |
| n_tp_escolaridade | 0,1,2,3,4,5,9 | Mother's education category | +110 |
| n_qt_nascidos_vivos | 0-18 | Number of live children | +120 |
| n_qt_nascidos_mortos | 0-19 | Number of deceased children | +140 |
| n_tp_gestacao | 0,1,2,3,4,5,6,9 | Weeks of gestation (by ranges) | +160 |
| n_tp_gravidez | 0,1,2,3,9 | Pregnancy type | +170 |
| n_tp_parto | 0,1,2,9 | Baby delivery type | +180 |
| n_tp_prenatal | 0,1,2,3,4,9 | Prenatal appointments (by ranges) | +190 |
| n_nu_apgar1 | 0-10 | Apgar score in the first minute | +200 |
| n_nu_apgar5 | 0-10 | Apgar score until fifth minute of live | +210 |
| n_nu_peso | 0-9999 | Weight in grams at birth | *0,03 |
| n_st_malformacao | 0,1,2,9 | Malformation at birth | +220 |
| n_tp_escolaridade_2010 | 0,1,2,3,4,5,9 | Mother's education category in 2010 | +230 |
| n_tp_raca_cor_mae | 0,1,2,3,4,5,9 | Mother's race and color category | +240 |
| n_qt_gestacao_anterior | 0-99 | Number of previous pregnancies | +130 |
| n_qt_parto_normal | 0-99 | Number of normal deliveries | n/a |
| n_qt_parto_cesarea | 0-99 | Number of cesarean deliveries | +100 |
| n_nu_semana_gestacao | 0-99 | Weeks of gestation | +200 |
| n_tp_metodo_estimar | 0,1,2,9 | Weeks of pregnancy estimation method | +240 |
| n_nu_mes_gestacao_prenatal | 0-10 | First prenatal appointment month | +10 |
| n_tp_apresentacao | 0,1,2,3,9 | Newborn presentation type | +20 |
| n_st_trabalho_parto | 0,1,2,3,9 | Induced labor? | +30 |
| n_st_cesarea_parto | 0,1,2,3,9 | Cesarean occurred before labor began? | +40 |
| n_tp_nascimento_assistido | 0,1,2,3,4,9 | Birth was attended? | +50 |
| n_tp_funcao_responsavel | 0,1,2,3,4,5 | Delivery responsible occupation | +60 |
| n_nu_idade_pai | 0-99 | Father's age | +70 |
| n_nu_serie_escolaridade_mae | 0-8 | Mother's basic school serie | +170 |
| n_co_uf_ibge_naturalidade | 0-99 | Mother's birth state code number | +180 |
| n_tp_grupo_robson | 0-11 | Robson Group code | +240 |
| n_tp_escolaridade_agregado1 | 0-12 | Mother's aggregated scholarly 1 | +25 |
| n_tp_escolaridade_agregado2 | 0-12 | Mother's aggregated scholarly 2 | +40 |
| month_birth | 0-12 | Month of birth | = 0 |
| year_birth | 2000-2016 | Year of birth | = 0 |
| month_death | 0-12 | Month of death | = 0 |

## 3 Proposed Method

The proposed method can be described in two main stages: as depicted on Figure 2, the first stage consists in building an image dataset by encoding the feature vector of each sample into a 6 × 6 gray scale image, using the methodology described in Section 3.1; and the second stage, classify using DemogNet, which consists in a new CNN architecture to classify image samples, produced on previous step, according to its classes, that is death (positive class) or alive (negative class).

**Fig. 2.**
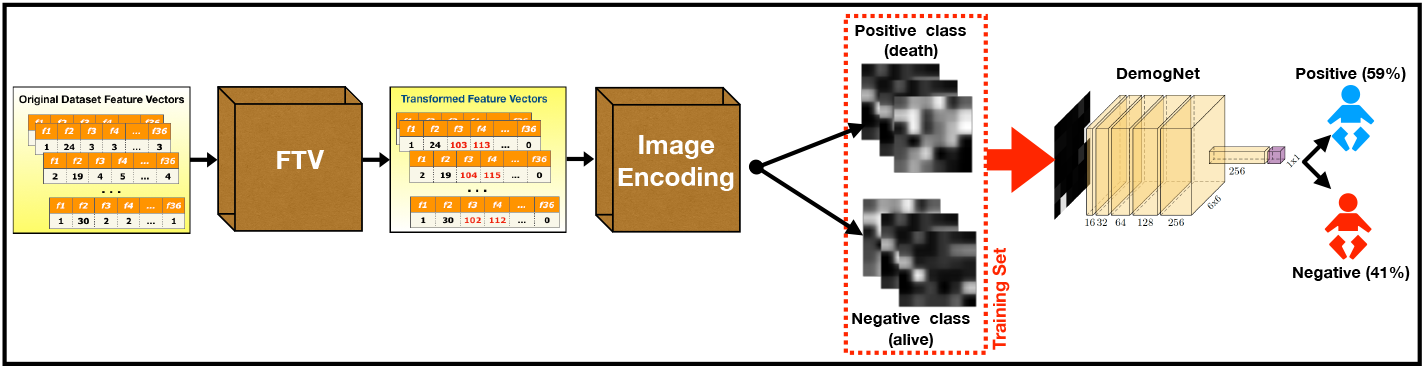
Overview of proposed method.

### 3.1 Stage #1: Encoding Categorical Data into Images

First step of proposed method consists in encoding feature vectors of each sample (which are composed by categorical features) into a grayscale image, in order to be inputted in a CNN.

Given that each sample is composed by a feature vector with 36 features, a 6 × 6 image have been created for each sample, where each pixel of the image refers to the value of one feature, converted to a gray scale value.

Furthermore, trying to improve classes separability, we proposed an additional approach, which was named as Feature Transformation Value (FTV) process. It consists in defining ranges within the total gray scale range (from 0 to 255), and associate an specific range to a specific feature, so features with identical original values will be encoded into different gray scale values.

Although FTV being a simple process, some specific features needed to be processed in specific ways. The features birth month, birth year and month death were truncated to zero due its irrelevance for the problem. Features birth place code, mother age and number of normal deliveries were left with its original value, just to follow the sequence of “range allocation”. The only exception is feature weight in grams at birth, which has its values multiplied by 0.03, in order to fit it into gray scale range. The FTV applied to each feature is defined on Table 1.

### 3.2 Stage #2: Demographic Network (DemogNet) for Classification

When encoded into images, our categorical data result in a 6 × 6 image. This kind of image resolution is a limitation for state-of-the-art CNN architectures, since most of them use as input square images with resolution between 200 and 300 pixels. Furthermore, images with low resolution make hard to adjust weights of very deep architectures, most of times leading to network instability and overfitting.

To deal with these kinds of particularities, we propose a new CNN architecture named Demographic Network (DemogNet). DemogNet uses a single convolutional block composed by five convolutional layers with an increment in the number of filters: 16, 32, 64, 128 and 256 respectively. Each convolutional layer uses 3 × 3 kernels. Activation on convolutional layers are performed using rectifier function (ReLU) [10]. In the end of convolutional block we also include a dropout of 0.25 to improve network generalization properties.

Next, we include a fully-connected layer, comprising 256 neurons with ReLU activation and dropout of 0.25.

Finally, a fully-connected layer comprising two neurons with a softmax activation [9] is included for classification purpose.

For optimization the method uses Stochastic Gradient Descendent (SGD) with a categorical cross-entropy as loss function [9].

## 4 Experiments and Results

This section presents details about experiments performed to validate proposed method, discussing obtained results.

### 4.1 Experimental Setup and Hyperparameters Choice

Proposed method have been implemented using Python (3.6) programming language and Scikit-Learn (0.21.2), Keras (2.2.4), Tensorflow (1.13.0), Pandas (0.24.2) and MatplotLib (3.1) as main libraries. All the experiments have been performed using a machine containing 40 CPU cores, 4 GPU TitanX 12 GB, 120 GB of RAM and 8 TB of storage, running Ubuntu 18.04 (64 bits).

DemogNet have been trained using SGD with a learning rate of 0.00001 and a momentum of 0.9 along 300 epochs.

### 4.2 Round #1: Balanced Dataset

In the first round of experiments we evaluate performance of proposed method in a subsample extracted from the main dataset. It contains the same number of samples for each class (death and alive).

Sub-samples have been extracted by selecting all images of positive (death) class (7,928 samples). Then, we randomly select the same number of images from negative (alive) class, resulting in a dataset with 15,856 samples.

For validation purpose, we applied a 10-folds cross validation protocol, and report results using average ROC curves and accuracy.

Figure 3 depicts average ROC curve as dark blue line and standard deviation calculated from 10-folds depicted as shaded region around darker curve. Average accuracy was 89.54% with an standard deviation of 0.01% and an AUC of 0.95.

**Fig. 3.**
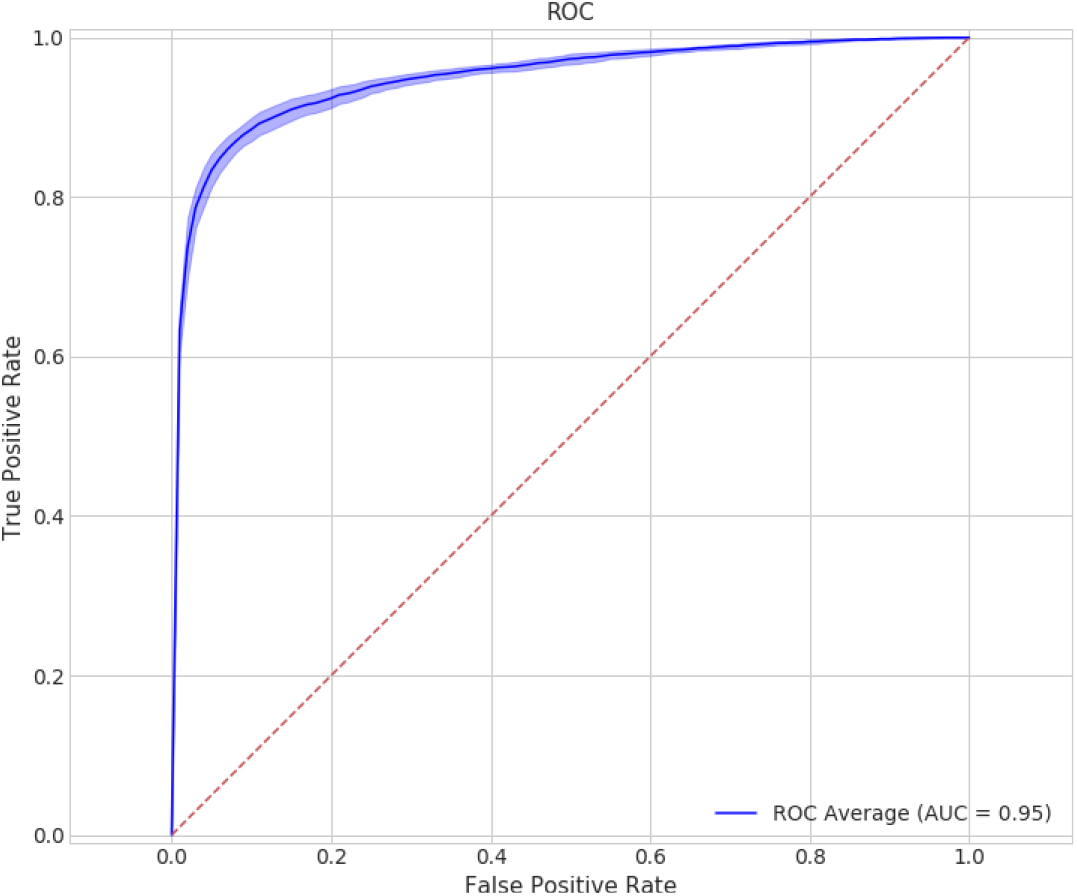
Average ROC and variance (shadow region) for 10-folds cross validation using balanced dataset.

Even using a small portion of the dataset, which results in a small number of samples when compared against other datasets as Imagenet [6], with a shallow CNN and working over gray scale images with small resolution (6 x 6), the proposed method presents good results, with a very small standard deviation across all 10 folds.

Train process converges after 200 epochs, becoming stable on loss decreasing and accuracy score increasing as depicted on Figures 4(a) and 4(b), respectively.

**Fig.4.**
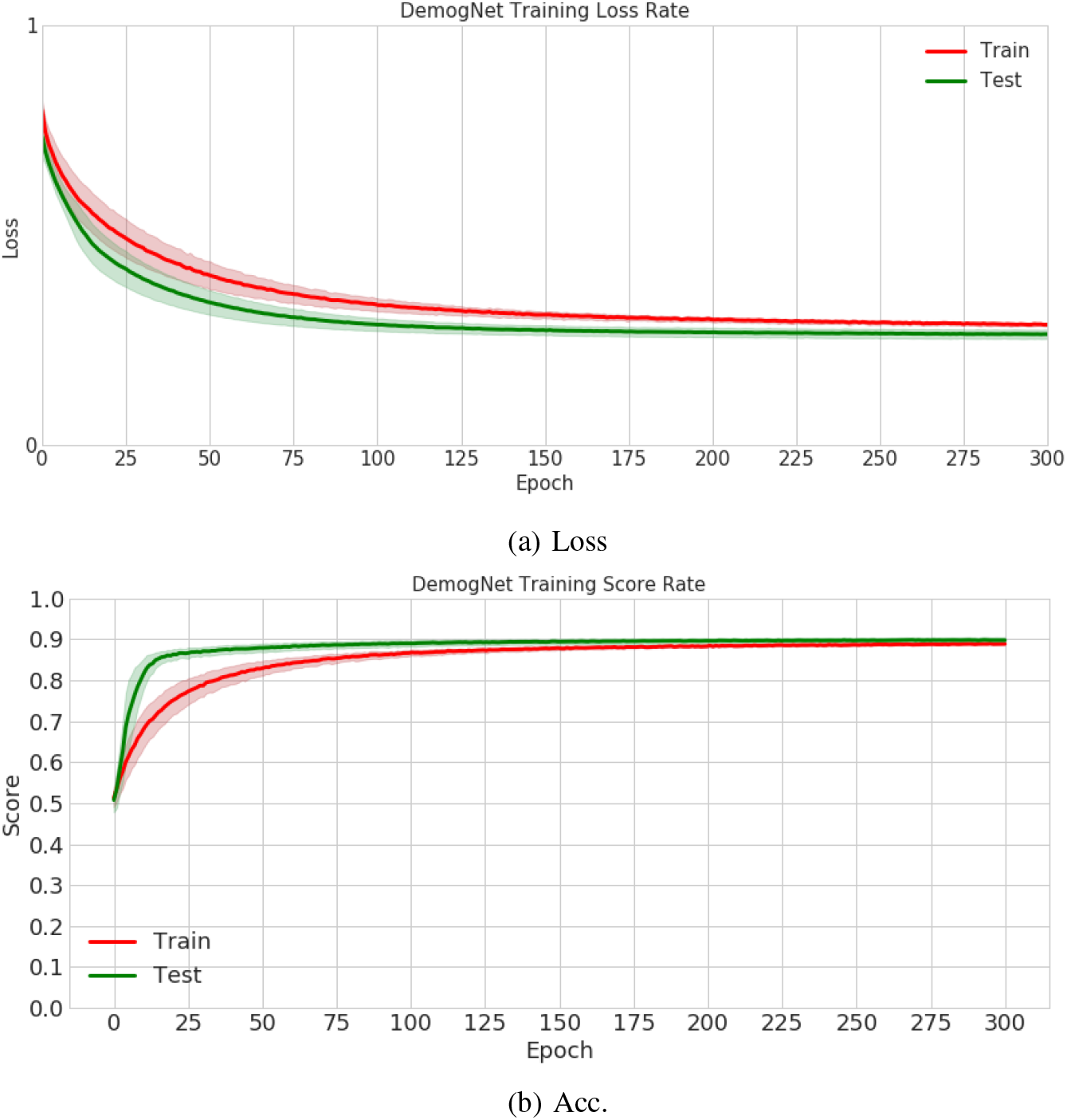
Loss and Accuracy scores on training process for balanced dataset.

### 4.3 Round #2: Unbalanced Dataset

Second round of experiments evaluates performance of proposed method when increasing the number of training samples and test proposed method using the entire dataset (excluding training samples).

For positive class (death), we randomly select 793 (10%) samples for test, 714(10%) samples for validation and remaining 6,421 samples composes the training set. In negative class (alive), on training and validation sets, for each positive sample we randomly select 5 negative samples, resulting in 32,105 and 3,570 respectively. Remaining negative samples have included test samples. This way, test set reflects real data distribution, which contains a massive number of negative samples against a small number of positive samples(1,392,232).

To compensate unbalanced data on training set, we use class weights, with weight of 1 for negative class and weight of 5 for positive class, on training process. It allows us to apply a higher penalty on misclassified positive samples. Figure 5 depicts ROC curve on this scenario.

**Fig. 5.**
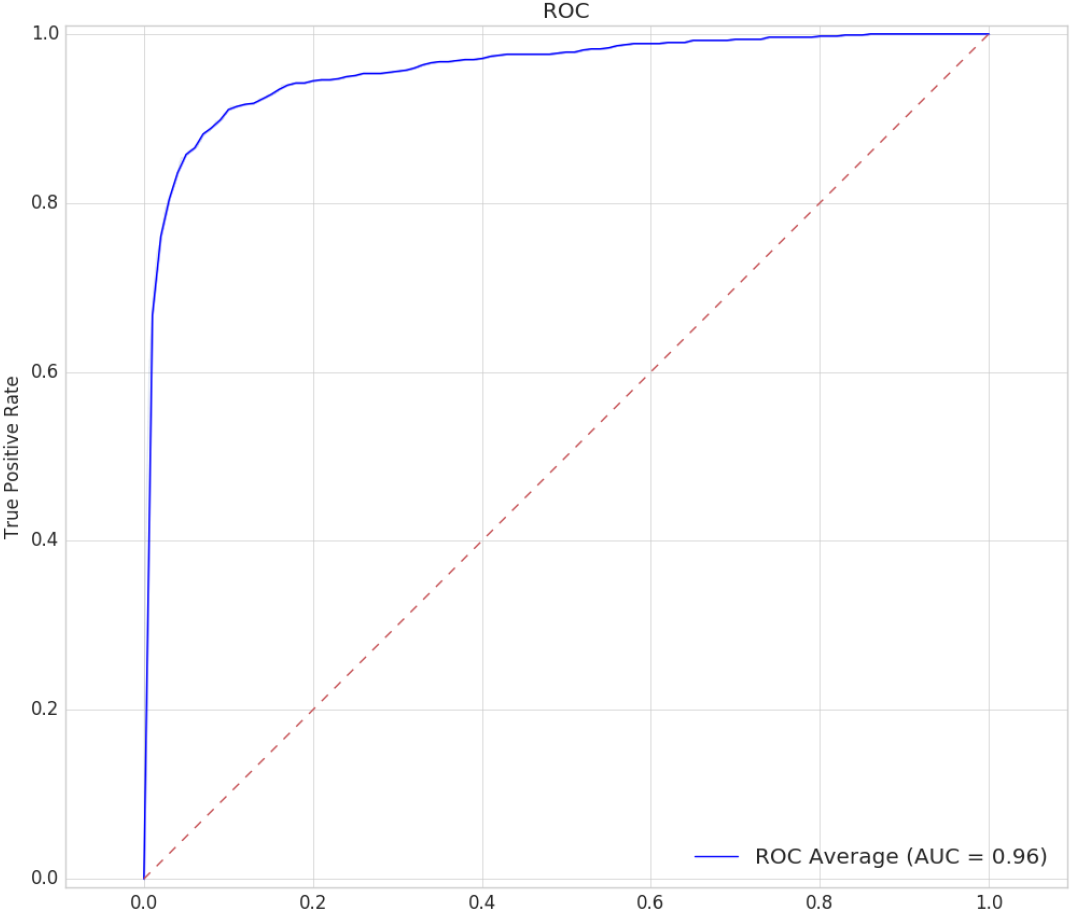
ROC curve for unbalanced dataset. This scenario reflects real data distribution, where positive (death) class represents less than 1% of dataset samples.

Accuracy achieved in the best ROC point was 90.61% with an AUC of 0.96. This result is very consistent with cross-validation results aforementioned, even when testing proposed method across more 1.4m samples. Reported results are very expressive and corroborate the claim of the method robustness.

As reported on Round #1, train process converges after 200 epochs, becoming stable on loss decreasing and accuracy score increasing as depicted on Figures 6(a) and 6(b), respectively. Small variations on loss of validation set can be associated with small number of validation samples, which can be easily corrected introducing more samples at the model.

**Fig.6.**
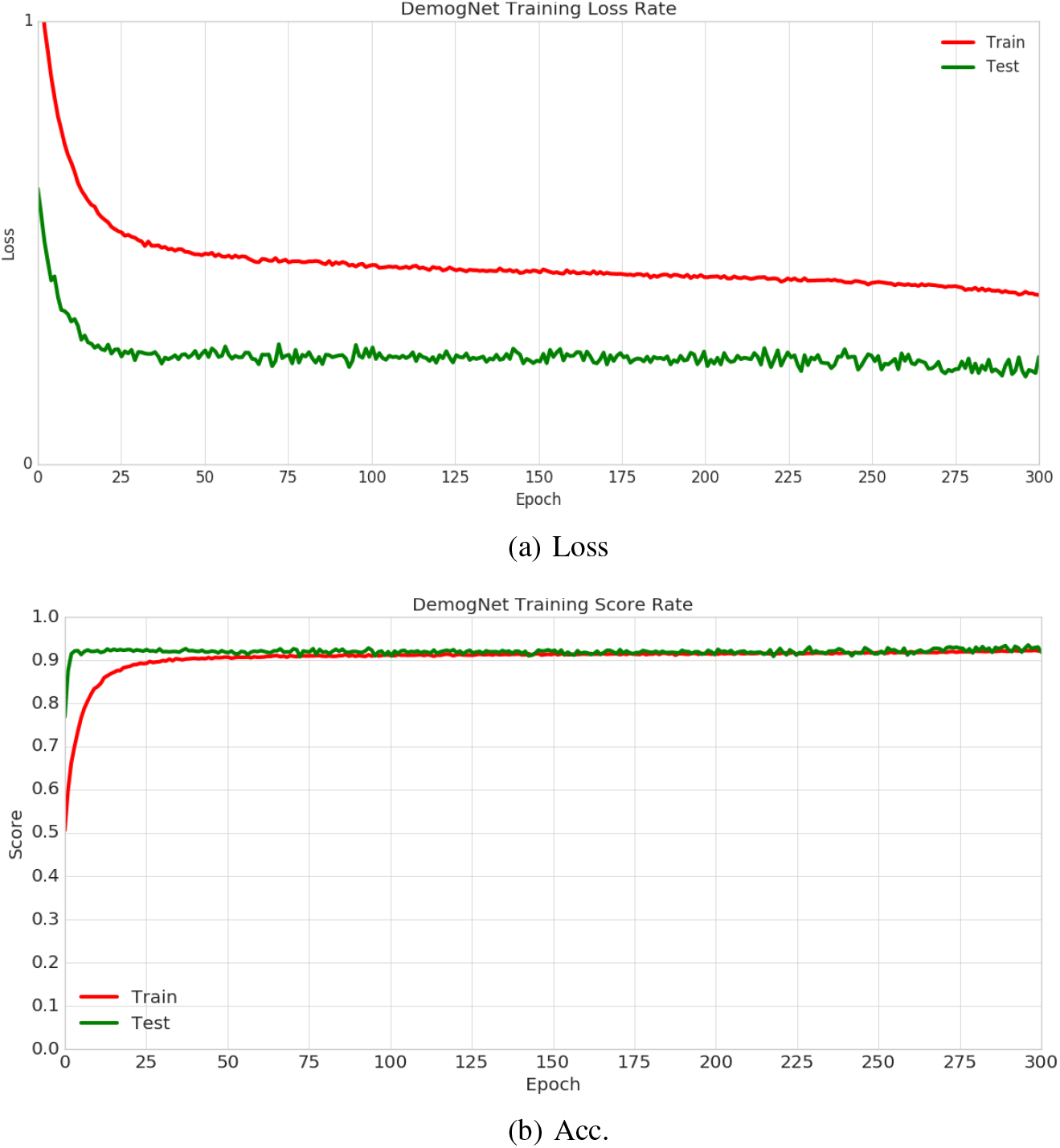
Loss and Accuracy scores on training process for unbalanced dataset.

### 4.4 Round #3: Comparison Against Standard Classifiers

Use of CNN to the problem of death risk classification based on demographic features is only justified if transformations applied over input image improves features in some way. Furthermore, to the best of our knowledge, there is no other work in literature based on similar features and data.

This round of experiments compares achieved results against standard classification methods, such as SVM (Support Vector Machines) and KNN (K-Nearest Neighbor) [5], in order to allow a benchmark for comparison with the proposed method. For SVM, we used an RBF kernel, along with a *qamma* of 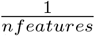 and a *C* value of 1.0. KNN algorithm was assessed with the number of neighbors set as 5.

Images by FVT are feed into classifiers (DemogNet, SVM and KNN) without any pre-processing and then classified. Table 2 present achieved results for balanced and unbalanced datasets.

**Table 2.** Comparison against standard classification methods

|  | ACC. (Balanced) | AUC. (Balanced) | ACC. (Unbalanced) | AUC. (Unbalanced) |
| --- | --- | --- | --- | --- |
| DemogNet | **0.89** | **0.95** | **0.91** | **0.96** |
| KNN | 0.83 | 0.90 | 0.84 | 0.90 |
| SVM | 0.58 | 0.91 | 0.58 | 0.91 |

DemogNet outperformed SVM and KNN in all scenarios which makes clear intermediate transformations performed by DemogNet contributes to increase separability

between classes.

### 4.5 Round #4: Providing Model Understanding

More than provide an answer about death risk, in methods designed to solve public health problems, interpretability is paramount. To address this problem, we take advantage of **SH**apley **A**dditive ex**P**lanation (SHAP) Values [15] method, which is able to highlight on images region where CNN is paying more attention in the moment of classification.

SHAP method belongs to “additive feature attribution methods” which can be simplified by linear function of features. SHAP tries to come up with a linear regression model for each data point. It replaces each feature (*x_i_*) with binary variable (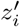) that represents whether *x_i_* is present or not into the model given by:

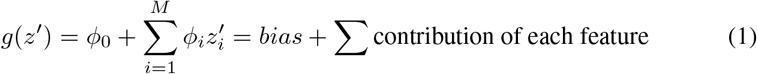

where *g(z′*) is a local surrogate model of the original linear model *f (x)* and *ϕ_i_* is how much the presence of feature *i* contributes to the final output, which helps to interpret the original model.

To measure the most important region (pixels) for each class, we calculated SHAP values for all test images, and then calculated the average map for positive and negative classes. Results are depicted on Figure 7

**Fig. 7.**
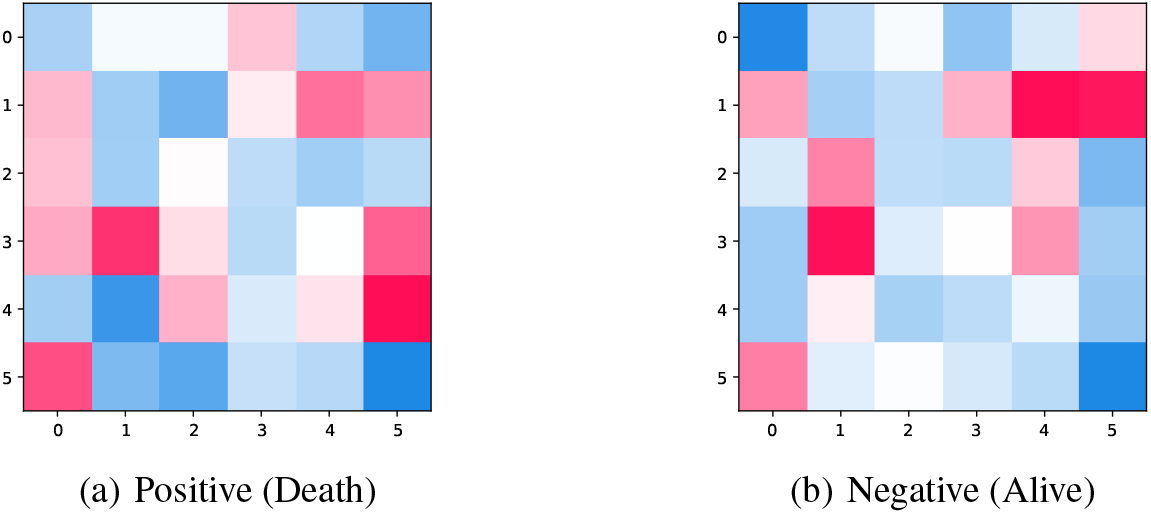
Average SHAP values indicating more important regions for positive and negative classes. As red value as features value, indicates a feature which influences model positively, while white and blue points indicates a feature which has almost no influence over model result.

As we can observe, positive and negative classes highlight different regions at images. While positive class are more influenced by a combination of features^4^ as weight on birth, number of live children, number of normal deliveries and induced labor, negative class is more influenced by features as occurence of malformation at birth, number of previous pregnancies and newborn presentation type.Both classes has an strong influence of apgar score until first and fifth minute of life [1] and weeks of pregnancy.

## 5 Conclusion and Future Works

This paper proposed a new method to address the problem neonatal death risk. Using data from Brazilian Information System of Live Births (SINASC) and from the Brazilian Information System of Mortality (SIM) of São Paulo city, we construct a dataset of death and alive records comprising more than 1.4m of samples with categorical features related with mother, pregnancy care, child features at born, etc. We then propose a new approach to encode this categorical data into small gray scale images.

Problem is modeled as a binary classification of death/alive classes problem and solved by a new CNN architecture, DemogNet, which is proper to deal with such small images at this specific case. Experiments performed along different setups show effectiveness of model classification, achieving an AUC of 0.96 in a dataset comprising more than 1.4M samples.

Furthermore, experiments demonstrate that application of a shallow CNN in classification outperform standard machine learning classification methods.

Additionally, performed experiments also help to better understand regions what contributes to CNN final answer, which is important for a better understanding of model results.

For future work, we intend to evaluate proposed method in a dataset to be constructed using data from all Brazilian states. Also, new methods for image construction from categorical data and the addition of new features meaningful to neonatal death risk will be evaluated.

## Data Availability

Data will be made public after paper acceptance in a journal.

## Footnotes

1 São Paulo is the capital city of the state of São Paulo in Brazil

2 On this study, we work using SIM and SINASC records just from São Paulo city.

3 CID10

4 Features at images are sorted according Table 1, which means, from top left to bottom right, first pixel represents first line on table (n_tp_ocorrencia), second pixel represents second line (n_nu_idade), and so on.

## Notes

### Competing Interest Statement

The authors have declared no competing interest.

### Funding Statement

This research was supported by Bill & Melinda Gates Foundation (Process no: OPP1201970) and Ministry of Health of Brazil, through the National Council for Scientific and Technological Development (CNPq) (Process no: 443774/2018-8). It was also supported by NVIDIA, that donated a GPU XP Titan used by the research team.

### Author Declarations

This paper uses publicly available data (SIM and SINASC) that has been de-identified and was deemed exempt from approval from a human research ethics committee.

## References

1. Apgar, V.: A proposal for a new method of evaluation of the newborn infant. Curr Res Anesth Analg 32, 260–267 (1953)

2. Arretche, M., Marques, E.: Municipalização da saúde no Brasil: diferenças regionais, poder do voto e estratégias de governo. Ciencia & Saúde Coletiva 7, 455–479 (00 2002)

3. Arruda, N.M.: Determinantes da Mortalidade Adulta nas Microrregiões Brasileiras em 2010: uma análise baseada em modelos de aprendizado de máquina. Master’s thesis, Universidade Estadual de Campinas, Campinas (2019)

4. Benhimane, S., Najafi, H., Matthias, G., Genc, Y., Navab, N., Malis, E.: Real-time object detection and tracking for industrial applications. In: VISAPP 2008: Proceedings of the Third International Conference on Computer Vision Theory and Applications (2008)

5. Bishop, C.M.: Pattern recognition and machine learning. springer (2006)

6. Deng, J., Dong, W., Socher, R., Li, L.J., Li, K., Fei-Fei, L.: ImageNet: A Large-Scale Hierarchical Image Database. In: CVPR09 (2009)

7. França, E., Lansky, S.: Mortalidade infantil neonatal no Brasil: situates, tendências e perspectivas. In: Rede Interagencial de Informação para a Saúde. Demografia e Saúde: contribuição para análise de situações e tendências. pp. 83–112. Organização Pan-Americana da Saúde(2009)

8. Garcia, L.P., Fernandes, C.M., Traebert, J.: Risk factors for neonatal death in the capital city with the lowest infant mortality rate in brazil. Jornal de Pediatria (Versão em Português) 95(2), 194–200 (2019). https://doi.org/10.1016/jjpedp.2018.03.004, https://linkinghub.elsevier.com/retrieve/pii/S225555361830034X

9. Goodfellow, I., Bengio, Y., Courville, A.: Deep Learning. MIT Press (2016), http://www.deeplearningbook.org

10. Hahnloser, R., Sarpeshkar, R., Mahowald, M.A., Douglas, R.J., Seung, H.S.: Digital selection and analogue amplification coexist in a cortex-inspired silicon circuit. Nature 405, 947–951(2000)

11. Hatami, N., Gavet, Y., Debayle, J.: Classification of time-series images using deep convolutional neural networks. In: Tenth International Conference on Machine Vision (ICMV 2017). vol. 10696 (2017)

12. Hsieh, M.H., Hsieh, M.J., Chen, C.M., Hsieh, C.C., Chao, C.M., Lai, C.C.: Comparison of machine learning models for the prediction of mortality of patients with unplanned extubation in intensive care units. Scientific Reports 8(1), 2045–2322 (2018)

13. Lansky, S., Friche, A.A.d.L., Silva, A.A.M.d., Campos, D., Bittencourt, S.D.d.A., Carvalho, M.L.d., Frias, P.G.d., Cavalcante, R.S., Cunha, A.J.L.A.d.: Pesquisa nascer no brasil: perfil da mortalidade neonatal e avaliação da assistência à gestante e ao recém-nascido. Cadernos de Saúde Pública 30, S192–S207 (00 2014)

14. Luan, S., Li, Y., Wang, X., Zhang, B.: Object detection and tracking benchmark in industry based on improved correlation filter. Multimedia Tools and Applications 77(22), 29919–29932 (Nov 2018)

15. Lundberg, S.M., Lee, S.I.: A unified approach to interpreting model predictions. In: Guyon, I., Luxburg, U.V., Bengio, S., Wallach, H., Fergus, R., Vishwanathan, S., Garnett, R. (eds.) Advances in Neural Information Processing Systems 30, pp. 4765-4774. Curran Associates, Inc. (2017)

16. Machado, J.C., Cotta, R.M.M., Soares, J.B.: Reflexões sobre o processo de municipalização das políticas de saúde: a questão da descontinuidade político-administrativa. Interface - Comunicação, Saúde, Educação 19, 159–170 (03 2015)

17. Mosley, W.H., Chen, L.C.: An analytical framework for the study of child survival in developing countries. 1984. Bulletin of the World Health Organization 81(2), 140–145 (2003), https://www.ncbi.nlm.nih.gov/pmc/articles/PMC2572391/

18. Pan, I., Nolan, L.B., Brown, R.R., Khan, R., v. d. Boor, P., Harris, D.G., Ghani, R.: Machine learning for social services: A study of prenatal case management in illinois. AJPH RESEARCH 107(6), 938–944 (2017). https://doi.org/10.2105/AJPH.2017.303711, http://ajph.aphapublications.org/doi/10.2105/AJPH.2017.303711

19. Podda, M., Bacciu, D., Micheli, A., Bellu, R., Placidi, G., Gagliardi, L.: A machine learning approach to estimating preterm infants survival: development of the preterm infants survival assessment (pisa) predictor. Scientific Reports 8(1), 2045–2322 (2018)

20. Rezende, E., Ruppert, G., Carvalho, T., Ramos, F., de Geus, P.: Malicious software classification using transfer learning of resnet-50 deep neural network. In: 2017 16th IEEE International Conference on Machine Learning and Applications (ICMLA). pp. 1011-1014 (Dec 2017). https://doi.org/10.1109/ICMLA.2017.00-19

21. Silva, A.L.A.d., Mendes, A.d.C.G., Miranda, G.M.D., Santos Neto, P.M.d.: Childbirth care in Brazil: a critical situation has not yet been overcome. 1999-2013. Revista Brasileira de Saude Materno Infantil 16, 129–137 (06 2016)

